# Omega-6 polyunsaturated fatty acids and adiposity in the UK Biobank Cohort: a cross-sectional and longitudinal prospective analysis

**DOI:** 10.1101/2025.09.24.25336554

**Authors:** Heidi T.M. Lai, Jason Westra, Evan De Jong, Nathan L. Tintle, Martha A. Belury, William S. Harris

## Abstract

**Background:** The role of omega-6 polyunsaturated fatty acids (PUFAs), especially linoleic acid (LA) in adiposity remains contested. While clinical interventions suggest improved body composition with higher LA intake, observational evidence is inconsistent, and few studies incorporate repeated measures to examine longitudinal change.

**Objective:** To investigate associations of circulating LA, non-LA omega-6, and total omega-6 PUFAs with adiposity outcomes in the UK Biobank.

**Methods:** Multivariable linear models evaluated cross-sectional and longitudinal associations between omega-6 fatty acid levels and waist circumference (WC), weight, and whole-body fat mass (FM) adjusting for relevant demographic, lifestyle and medical history covariates. Models considered FA levels per interquintile range and by quintiles.

**Results:** Cross-sectionally, higher circulating LA was inversely associated with WC, weight, and FM. Participants in the highest versus lowest quintile of LA had significantly smaller WC [–11.04 (–11.17, –10.91) cm], lower weight [–11.77 (–11.92, –11.62) kg], and lower FM [–7.87 (–7.97, –7.77) kg]. Associations for total omega-6 were generally consistent with those for LA. Conversely, non-LA omega-6 was positively associated with WC [1.46 (1.32, 1.61) cm], weight [2.41 (2.25, 2.58) kg], and FM [1.81 (1.69, 1.92) kg]. Longitudinal analyses largely corroborated these patterns, with annual changes in WC, weight, and FM inversely associated with LA and positively associated with non-LA omega-6.

**Conclusions:** Higher circulating LA, but not non-LA omega-6, was associated with lower WC, weight, and FM both cross-sectionally and longitudinally. Our findings support dietary recommendations to promote LA-rich oils. Divergent associations between LA and non-LA omega-6 caution against treating omega-6 PUFAs as a homogenous group, and there remains a need to examine the distinct health effects of individual non-LA omega-6.

## Introduction

Obesity affects 890 million adults worldwide [1] and is a major risk factor for cardiometabolic disease [2], contributing to more than 1.6 million premature deaths each year globally [3]. By 2035, the global economic burden attributable to obesity is projected to exceed $4 trillion annually (USD) [4]. As suboptimal diet is a modifiable risk factor for obesity [5], identifying determinants of adiposity is an urgent public health priority.

Linoleic acid (LA), an essential omega-6 polyunsaturated fatty acid (PUFA), accounting for 85-90% of omega-6 PUFA intake in the Western diet, and abundantly sourced from seed oils (e.g., soybean, corn, cottonseed oils), nuts, and seeds [6, 7]. The majority of non-LA omega-6 PUFAs in the diet and the blood consist of arachidonic acid (AA) [6]. Others such as dihomo-γ-linolenic acid (DGLA), γ-linolenic acid (GLA), adrenic acid, and osbond acid are predominantly determined by metabolic processes [7]. Although LA is well studied for its cardiometabolic effects [8], its role in body weight regulation remains contested [9].

Several arguments have been raised which point to potential adverse effects of all omega-6 PUFAs, or LA in particular. First, ecological observations show that rising omega-6 PUFAs levels in the diet (and a parallel decline in saturated fats) coincided with increased cardiometabolic disease rates in the early 20^th^ century [10]. Second, LA serves as a precursor to AA, which is a substrate for several pro-inflammatory eicosanoids [11], fueling speculation that a high LA intake may promote chronic systemic inflammation and obesity via its conversion to AA [7]. More recently, all omega-6 PUFAs (including LA) attracted adverse publicity related to its association with industrial food processing [12, 13]. In terms of mechanistic concerns, studies in animal models (C57BL/6j mice) demonstrated that elevated levels of dietary LA increased endocannabinoid production, promoted weight gain, and impaired insulin signalling [14, 15]. Specifically, a diet comprised of 22.5% energy intake from LA induced greater weight gain than saturated fat diets, despite the absence of hypothalamic inflammation [15]. However, this dose far exceeds typical human intakes, and the applicability of these findings to humans remain questionable.

In contrast, clinical interventions generally report reduced systemic inflammation and improved body composition with the addition of LA to the diet [9, 16–19], especially in lean mass [17, 19].

Observational cohorts also link higher levels of circulating total omega-6 PUFAs, LA and, in some cases, AA, to anti-inflammatory profiles [20–22]. In studies incorporating anthropometric measures, circulating LA was positively associated with lean tissue volume [23] and skeletal muscle mass [24], inversely associated with trunk adipose mass [23] and abdominal obesity [25], although associations for weight gain and body mass index are mixed [23, 26, 27]. Findings are less consistent for the associations between AA and BMI [26, 28], while other, non-LA omega-6 PUFAs have not been individually studied. LA was also inversely associated with metabolic syndrome [29, 30], suggesting that at population relevant exposures, higher LA is unlikely to promote, and might even attenuate weight gain.

Despite this body of work, pronounced gaps remain. Findings from studies which rely on dietary self-report methods are less consistent, reporting largely null associations between omega-6 overall or LA in particular with anthropometric measurements [28, 31–34], potentially reflecting exposure misclassification. Furthermore, few studies have incorporated repeated measures to assess baseline levels and longitudinal changes over time. Large-scale studies using objective biomarkers of LA status, particularly in relation to adiposity outcomes, are also sparse. To address this gap, we examined the cross-sectional and longitudinal relationship between circulating LA levels in relation to body weight and adiposity outcomes in the UK Biobank (UKBB), a large, prospective cohort.

## Methods

### Study population

The UKBB is a prospective, population-based cohort of ~500,000 individuals recruited between 2007 and 2010 at assessment centers across England, Wales, and Scotland. Baseline data was collected using questionnaires, biological samples, and physical measurements. Ongoing longitudinal monitoring occurs via a mix of in-person measurements, in-person and online questionnaires, as well as nearly real-time electronic medical record and death registry integration [35, 36]. UKBB has ethical approval (Ref. 11/NW/0382) from the North West Multicentre Research Ethics Committee. Use of these de-identified, publicly available data for research was approved by the University of South Dakota Institutional Review Board (IRB-21-147). All participants provided electronic signed informed consent. The UKBB study was conducted according to the guidelines laid down in the Declaration of Helsinki. The UKBB protocol is available online (http://www.ukbiobank.ac.uk/wp-ontent/uploads/2011/11/UK-Biobank-Protocol.pdf). To be included, participants needed to have data on FAs and all covariates used in the analysis. Of 502,128 subjects in the UKBB, FA data were available on a random sample of 274,003. After removing 1,416 individuals with incomplete information on covariates (anthropometric measures, N=1090; Townsend Deprivation Index [37], N=326), the final sample available for cross-sectional analysis was 272,587 (Supplemental Figure 1). For longitudinal associations, we used the subsample of individuals who attended the repeat assessment (N = 20,295), the imaging visit (N = 88,641) or the first repeat imaging visit (N=12,591) [38]. Of these 99,044 unique individuals, a random sample of 59,394 had available fatty acid data, and 1,059 more were removed for missing covariates, resulting in 58,335 participants.

### Exposure

Our two primary exposures are plasma LA and non-LA omega-6 (each expressed as a percentage of total plasma FAs). The secondary exposure is total omega-6. The level of total omega-6 and LA was determined on baseline plasma samples using nuclear magnetic resonance (NMR; Nightingale Health) [39]; analytical performance characteristics have been reported [40]. Non-LA omega-6 is computed as the difference between total omega-6 and LA.

### Outcomes

Our primary outcomes are waist circumference, weight, and whole-body fat mass. Secondary outcomes included body mass index (BMI), whole-body fat-free mass, trunk fat mass, and trunk fat percent. For cross-sectional associations, we relied on anthropometric measurements taken at baseline (2006-10). For analyses that explored changes over time, we relied on follow-up measurements taken in 2012-13 during the first repeat assessment, 2014 and onwards for an imaging visit, and 2019 and onwards for the first repeat imaging visit. Waist circumference was taken by a Seca 200 cm tape measure, standing height by Seca 240 cm height measure, and the remaining measurements were collected by a Tanita BC418MA body composition analyser, all via standard protocol (https://biobank.ctsu.ox.ac.uk/ukb/ukb/docs/Anthropometry.pdf).

### Covariates

Information on all reported sociodemographic characteristics, diet, and lifestyle factors were collected via a touchscreen questionnaire. The electronic questionnaire and other resources can be found on the UKBB website (http://www.ukbiobank.ac.uk/resources/). Details of covariates used and their corresponding ID in the UKBB database can be found in Supplementary Table 1.

### Statistical analysis

Sample characteristics were summarized using standard approaches (mean/SD; N/%). The associations between the exposures of interest (LA and non-LA omega-6 PUFAs) and cross-sectional outcome measures were assessed with multivariable linear models, that adjusted for relevant covariates. Longitudinal models predicted changes in outcome measures divided by the length of time (years) between measurements to account for person-to-person differences in exposure time, and adjusting for relevant covariates. Associations are expressed as change in the outcome measure per interquintile range (IQ_5_R, defined as 90^th^ minus 10^th^ percentiles of each exposure of interest), or per quintile (Q; relative to the lowest quintile [Q1]) and via a linear trend across quintiles. Exploratory analyses used restricted cubic splines to test for potential non-linearity vs. linear models between LA and the three primary cross-sectional and three primary longitudinal outcomes. Interaction terms between continuous LA (IQ_5_R) and sex or (separately) age decade (40-50, 50-60, 60-70) were added to each model (cross-sectional and longitudinal) to test for potential sex or age modification of the LA– outcome relationships. A significance level of 0.05 was used for all analyses.

## Results

### Participant characteristics

We evaluated a total of 272,587 participants for the cross-sectional analysis (**Table 1**), and 58,335 participants for the longitudinal analysis (**Table 2**). Distributions of sex, race, education, Townsend Deprivation Indices, and physical activity were similar between the cross-sectional and longitudinal sub-cohorts. About half were female, nearly all were White, with just about half reporting a college education. Participants also tended to be slightly overweight at baseline, with a general reduction in weight and whole-body fat free mass during follow-up. Correlations between anthropometric measures are reported in Supplementary Table 2.

**Table 1:** Participant characteristics of the UK Biobank (cross-sectional analysis)

| **Variable** | **Mean (SD) or Frequency (%)** |
| --- | --- |
| Sex |  |
| Female | 147,176 (54) |
| Male | 125,411 (46) |
| Age (years) | 56.6 (8.08) |
| Race |  |
| White | 258,175 (94.7) |
| Asian | 5,627 (2.1) |
| Black | 3,807 (1.4) |
| Missing | 1,205 (0.4) |
| Other | 3,773 (1.4) |
| Education |  |
| College | 145,529 (53.4) |
| High School | 76,191 (28.0) |
| Less Than High School | 47,735 (17.5) |
| Missing | 3,132 (1.1) |
| Townsend score | -1.36 (3.07) |
| Physical Activity |  |
| Lowest | 60,661 (22.3) |
| Low | 62,012 (22.7) |
| High | 62,299 (22.9) |
| Highest | 63,192 (23.2) |
| Missing | 24,423 (9.0) |
| *Fatty acid measurements (% total fatty acid)* |  |
| Linoleic acid (LA) | 28.9 (3.44) |
| Omega-6 PUFA | 37.9 (3.63) |
| Omega-6: non-LA | 9.0 (1.91) |
| Docosahexaenoic acid (DHA) | 2.0 (0.68) |
| Omega-3 PUFA | 4.4 (1.55) |
| Omega-3: non-DHA | 2.4 (1.01) |
| *Anthropometric measures* |  |
| Standing Height (cm) | 168.5 (9.26) |
| Weight (kg) | 78.1 (15.89) |
| BMI (kg/m<sup>2</sup>) | 27.5 (4.78) |
| Waist Circumference (cm) | 90.3 (13.45) |
| Whole-body Fat Free Mass (kg) | 53.3 (11.49) |
| Whole-body Fat Mass (kg) | 24.9 (9.53) |
| Trunk Fat Mass (kg) | 13.8 (5.16) |
| Trunk Percent Fat (%) | 31.2 (8.0) |

**Table 2:** Participant characteristics of the UK Biobank (longitudinal analysis)

| **Variable** | **Mean (SD) or Frequency (%)** |
| --- | --- |
| Sex |  |
| Female | 30,198 (51.8) |
| Male | 28,137 (48.2) |
| Age (years) | 55.4 (7.63) |
| Race |  |
| White | 56,425 (96.7) |
| Asian | 765 (1.3) |
| Black | 374 (0.6) |
| Missing | 164 (0.3) |
| Other | 607 (1.0) |
| Education |  |
| College | 37,403 (64.1) |
| High School | 16,265 (27.9) |
| Less Than High School | 4,434 (7.6) |
| Missing | 233 (0.4) |
| Townsend score | -1.93 (2.73) |
| Physical Activity |  |
| Lowest | 13,154 (22.5) |
| Low | 14,673 (22.2) |
| High | 15,071 (25.8) |
| Highest | 12,924 (22.2) |
| Missing | 2,513 (4.3) |
| *Fatty acid measurements (% total fatty acid)* |  |
| Linoleic acid (LA) | 29.3 (3.33) |
| Omega-6 PUFA | 38.2 (3.5) |
| Omega-6: non-LA | 8.9 (1.86) |
| Docosahexaenoic acid (DHA) | 2.1 (0.67) |
| Omega-3 PUFA | 4.5 (1.55) |
| Omega-3: non-DHA | 2.4 (1.01) |
| *Anthropometric measures (baseline)* |  |
| Standing Height (cm) | 169.6 (9.15) |
| Weight (kg) | 77.2 (15.06) |
| BMI (kg/m<sup>2</sup>) | 26.8 (4.34) |
| Waist Circumference (cm) | 88.4 (12.77) |
| Whole-body Fat Free Mass (kg) | 53.7 (11.41) |
| Whole-body Fat Mass (kg) | 23.5 (8.8) |
| Trunk Fat Mass (kg) | 13.1 (4.84) |
| Trunk Percent Fat (%) | 30.0 (7.4) |
| *Anthropometric measures (change per year) <sup>1</sup>* |  |
| Weight (kg) | -0.08 (0.71) |
| BMI (kg/m<sup>2</sup>) | 0.00 (0.25) |
| Waist Circumference (cm) | 0.14 (0.94) |
| Whole-body Fat Free Mass (kg) | -0.16 (0.36) |
| Whole-body Fat Mass (kg) | 0.08 (0.64) |
| Trunk Fat Mass (kg) | 0.05 (0.39) |
| Trunk Percent Fat (%) | 0.11 (0.65) |

<sup>1</sup> Paired t-tests were conducted between baseline and repeated measures: all were statistically significantly different (p<0.001), except for BMI (p=0.135).

### Cross-sectional relationship between levels of omega-6 and primary measures

In multivariable models, LA was inversely associated with waist circumference, weight, and whole-body fat mass (**Figure 1**). In comparison to the lowest quintile, participants in the highest quintile had a statistically significant smaller waist circumference [−11.04 (−11.17, −10.91) cm], weighed less [−11.77 (−11.92, −11.62) kg], and had less whole-body fat mass [−7.87 (−7.97, −7.77) kg] (Supplementary Table 2). While findings for total omega-6 echoed those of LA (all p<0.001) (Supplementary Table 3), non-LA exhibited statistically significant associations in the opposite direction (**Figure 1**), though the magnitude of the relationships were smaller. Participants in the highest quintile had a slightly higher waist circumference [1.46 (1.32, 1.61) cm], were heavier [2.41 (2.25, 2.58) kg], and had more whole-body fat mass [1.81 (1.69, 1.92) kg] compared to the lowest quintile (Supplementary Table 3). All associations remained robust and statistically significant when fatty acid levels were assessed continuously per IQ_5_R (Supplementary Table 3).

**Figure 1:**
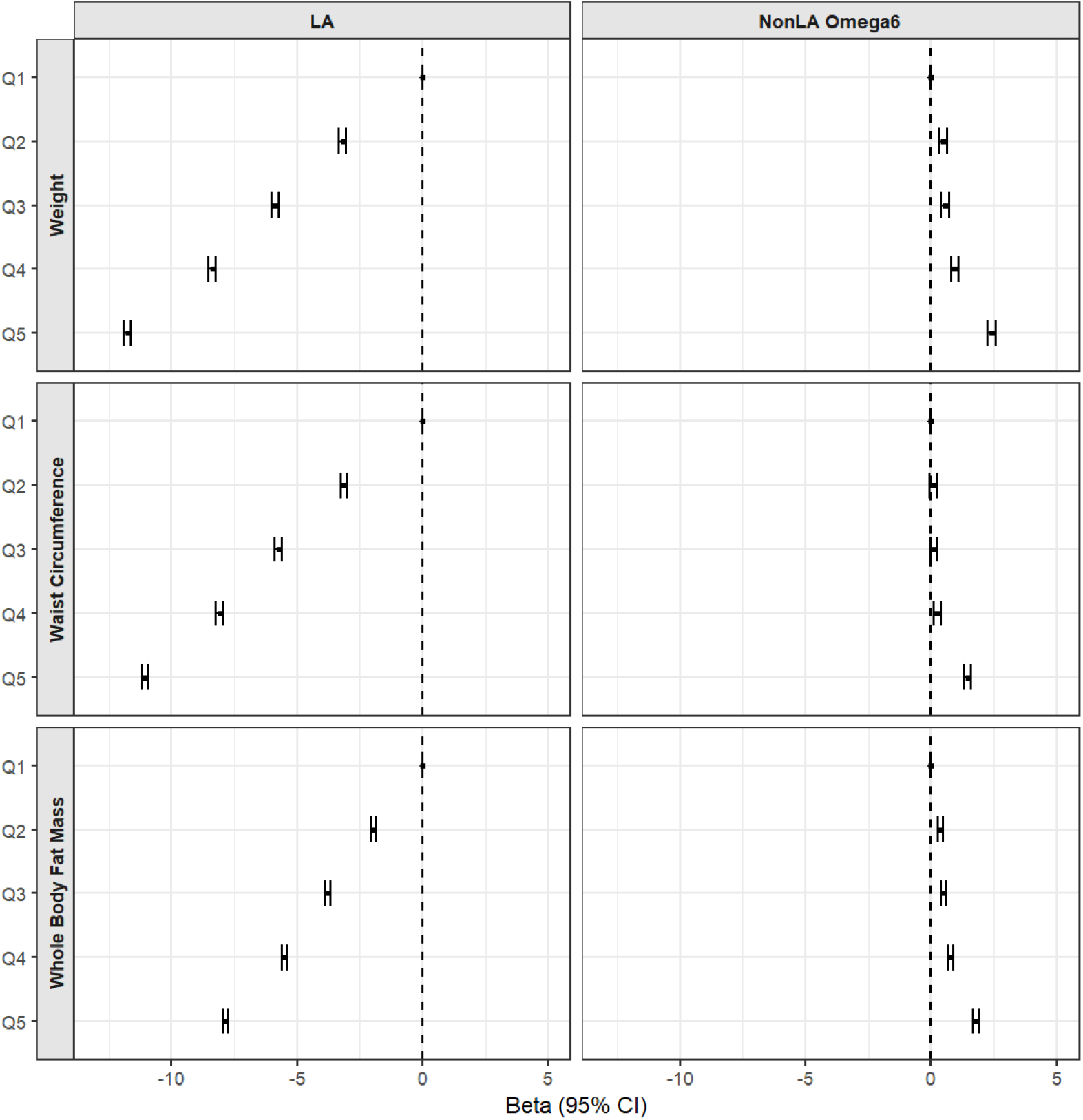
The cross-sectional association between linoleic acid (% total fatty acids) and weight (kg), waist circumference (cm), and whole-body fat mass (kg) in the UK Biobank.

### Changes in primary anthropometric measures over time

Similar to the results from our cross-sectional analysis, anthropometric measures decreased over time with higher levels of LA (**Table 3**). Waist circumference, weight, and whole-body fat mass for participants in the highest quintile decreased by −0.11 (−0.13, −0.09) cm, −0.05 (−0.07, −0.03) kg, and −0.04 (−0.06, −0.02) kg per year, respectively. Similarly, rates of change per year by IQ_5_R followed the results of analyses by quintiles, showing a statistically significant decrease in all three outcomes. Results for total omega-6 remain followed those of LA (Supplementary Table 4). For non-LA omega-6, waist circumference, weight, and whole-body fat mass increased slightly over time. Extreme-quintile differences are an increase of 0.09 (0.07, 0.12) cm, 0.13 (0.11, 0.14) kg, and 0.12 (0.10, 0.14) kg per year, respectively: in line with findings per IQ_5_R (Table 4).

**Table 3:** The longitudinal association between linoleic acid and change in weight, waist circumference, and whole-body fat mass in the UK Biobank.

|  | Quintiles of fatty acid |  |  |  |  |  |  |  |
| --- | --- | --- | --- | --- | --- | --- | --- | --- |
| *Linoleic acid* | I | II | III | IV | V | p- trend<sup>1</sup> | Per IQ5R<sup>2</sup> | p-value |
| Total N | 11,654 | 11,657 | 11,689 | 11,679 | 11,656 |  | 58,335 |  |
| Median (% total fatty acid) | 25.0 | 27.8 | 29.5 | 31.1 | 33.3 |  | 29.5 |  |
| Waist circumference (cm) <sup>3</sup> | 0 (Ref) | -0.01 (-0.04, 0.01) | -0.04 (-0.07, -0.02) | -0.07 (-0.10, -0.05) | -0.11 (-0.13, -0.09) | <0.001 | -0.10 (-0.12, -0.08) | <0.001 |
| Total N | 11,654 | 11,657 | 11,689 | 11,679 | 11,656 |  | 58,335 |  |
| Median (% total fatty acid) | 25.0 | 27.8 | 29.5 | 31.1 | 33.3 |  | 29.5 |  |
| Weight (kg) <sup>3</sup> | 0 (Ref) | 0.02 (0.00, 0.04) | 0.01 (-0.01, 0.03) | 0.00 (-0.02, 0.01) | -0.05 (-0.07, -0.03) | <0.001 | -0.03 (-0.05, -0.02) | <0.001 |
| Total N | 8,875 | 8,805 | 8,615 | 8,504 | 8,306 |  | 43,105 |  |
| Median (% total fatty acid) | 25.0 | 27.7 | 29.5 | 31.1 | 33.3 |  | 29.4 |  |
| Whole-body fat mass (kg) <sup>3</sup> | 0 (Ref) | 0.01 (0.00, 0.03) | 0.00 (-0.02, 0.02) | -0.01 (-0.03, 0.01) | -0.04 (-0.06, -0.02) | <0.001 | -0.03 (-0.05, -0.02) | <0.001 |
| *Non-LA omega-6* |  |  |  |  |  |  |  |  |
| Total N | 11,668 | 11,692 | 11,667 | 11,665 | 11,643 |  | 58,335 |  |
| Median (% total fatty acid) | 6.6 | 7.9 | 8.8 | 9.8 | 11.3 |  | 8.8 |  |
| Waist circumference (cm) <sup>3</sup> | 0 (Ref) | 0.02 (0.00, 0.04) | 0.03 (0.01, 0.06) | 0.03 (0.01, 0.05) | 0.09 (0.07, 0.12) | <0.001 | 0.09 (0.07, 0.11) | <0.001 |
| Total N | 11,668 | 11,692 | 11,667 | 11,665 | 11,643 |  | 58,335 |  |
| Median (% total fatty acid) | 6.6 | 7.9 | 8.8 | 9.8 | 11.3 |  | 8.8 |  |
| Weight (kg) <sup>3</sup> | 0 (Ref) | 0.03 (0.01, 0.05) | 0.05 (0.04, 0.07) | 0.06 (0.05, 0.08) | 0.13 (0.11, 0.14) | <0.001 | 0.12 (0.10, 0.13) | <0.001 |
| Total N | 8,543 | 8,602 | 8,536 | 8,613 | 8,811 |  | 43,105 |  |
| Median (% total fatty acid) | 6.7 | 7.9 | 8.8 | 9.8 | 11.3 |  | 8.8 |  |
| Whole-body fat mass (kg) <sup>3</sup> | 0 (Ref) | 0.03 (0.01, 0.05) | 0.06 (0.04, 0.08) | 0.07 (0.05, 0.09) | 0.12 (0.10, 0.14) | <0.001 | 0.11 (0.10, 0.13) | <0.001 |

Abbreviations: Linoleic acid, LA; inter-quintile range, IQ5R
Models are adjusted for age, sex, race, standing height, baseline weight, baseline waist circumference, baseline whole body fat mass, education, Townsend scores, physical activity, and levels of DHA.
<sup>1</sup> P-trend is generated by assigning participants the median value in each quintile then assessing quintiles as continuous variables
<sup>2</sup> Interquintile range (IQ5R) is the difference between the first and fifth quintiles
<sup>3</sup> Findings are reported as the difference in comparison to the lowest quintile in terms of change per year. Negative values mean that the outcome of interest is smaller compared to the reference group (set as zero), while positive values indicate the opposite.

### Associations between omega-6 levels and secondary outcomes

Cross-sectional associations with secondary outcomes (BMI, whole-body fat-free mass, trunk fat mass, and trunk fat percent) are reported in Supplementary Table 5. In brief, the direction of associations aligned with the primary outcomes, i.e., a higher level of LA and total omega-6 was linked to a lower BMI, whole-body fat-free mass, trunk fat mass, and trunk fat percent, while non-LA omega 6 demonstrated a statistically significant relationship in the opposite direction. For longitudinal associations, minor differences were present in comparison to primary outcomes (Supplementary Table 6). For example, the association between LA and whole-body fat-free mass was neutral, and total omega-6 was linked to increased levels of whole-body fat-free mass, but not trunk mass or trunk percent fat.

### Evidence of non-linearity

Restricted cubic splines showed evidence of non-linearity for the cross-sectional relationships between LA and all three primary outcomes (all three *p<0*.*001*). At lower LA values (20% to 25% of total fatty acids), associations showed a weaker inverse association with waist circumference, weight, and whole-body fat mass (Supplementary Figures 1 to 3) which strengthened as LA levels increased beyond 25%, though it weakened again beyond 35%. Similarly, all longitudinal models showed evidence of significant non-linearity (all three p<0.05). Specifically, LA had the strongest relationships with yearly changes in waist circumference, weight, and whole-body fat mass (Supplementary Figures 4 to 6) at levels of LA beyond 32%, with less evidence of a relationship for levels of LA levels less than 29%.

### Interactions with age and sex

Cross-sectionally, the strength of the inverse relationship between LA waist circumference, weight, and whole-body fat mass were statistically significantly stronger (all five *p <0*.*001*) among participants who were younger or female (Supplementary Table 7), in comparison to older or male participants, respectively. In longitudinal analyses, only weight change (with age; p<0.05) and waist circumference change (with sex; p<0.001) had statistically significant interactions. The stronger inverse relationship between LA and waist circumference was only present in participants who were female. Similarly, the stronger inverse association between LA and body weight was only present in those who were younger (Supplementary Table 8).

## Discussion

In our prospective cohort of over a quarter million participants, cross-sectional analyses showed that LA and total omega-6 PUFAs were inversely associated with waist circumference, weight, and whole-body fat mass. By contrast, higher levels of non-LA omega-6 were linked to greater waist circumference, weight, and whole-body fat mass, though the absolute values were small. Longitudinal analyses assessing annual change largely corroborated the cross-sectional findings. Findings for secondary outcomes (BMI, whole-body fat-free mass, trunk fat mass, and trunk fat percent) are also aligned with primary outcomes. We also observed evidence for non-linearity in the associations between LA and waist circumference, weight, and whole-body fat mass. All associations remain inverse and statistically significant after accounting for interactions with age and sex, although the strength of associations varied across subgroups.

Our findings show that circulating levels of LA are inversely associated with waist circumference, weight, and whole-body fat mass, supporting its potential protective role in body-weight regulation. Randomized controlled trials reinforce this biologic plausibility, demonstrating that LA may favourably influence adiposity through mechanisms such as improved insulin resistance [16, 23], maintenance of lean tissue [16, 23], reducing visceral fat [17, 41], and attenuating chronic inflammation [23, 42, 43]. Dietary fortification of LA directly increases the level of LA in the blood and tissue [44]. Mechanistically, LA and its oxylipin metabolites act on G protein–coupled receptors and peroxisome proliferator–activated receptors (PPAR) (PPARα, PPARβ/δ, PPARγ), modulating downstream pathways governing energy production and utilization [9]. In addition, through the cytochrome P450 pathway, LA-derived vicinal diols [9,10-dihydroxy-9Z-octadecenoic acid (DiHOME), 12,13-DiHOME] are inversely associated with adiposity [45, 46], while levels of LA-derived epoxides [9(10)-epoxyoctadecenoic acid (EpOME), 12(13)-EpOME] are lower in subjects with metabolic syndrome [47].

In contrast, non-LA omega-6 PUFAs were positively associated with waist circumference, weight, and whole-body fat mass, albeit with small effect sizes. Interpretation is challenging, as the non-LA omega-6 fraction consists of several different omega-6 PUFAs, including the predominant AA, as well as DGLA, GLA, adrenic acid, and osbond acids. While AA is known to exhibit pro-inflammatory effects, some of its metabolites (prostaglandin E2), are also inflammation resolvers (inhibition of tumour necrosis factor-α; inducing production of lipoxin A4) [7]. Similarly, oxylipins derived from other non-LA omega-6, e.g., DGLA, may exert effects distinct from those of AA [48]. Hence, the findings on non-LA omega-6s provide limited insight because the individual fatty acid associations cannot be evaluated.

Nevertheless, several takeaways can be drawn. If we consider findings for total omega-6, the overall association is inverse, suggesting that the strong inverse association with LA offsets the weak positive association of non-LA omega PUFAs, resulting in an overall benefit. The divergence between LA and non-LA omega fractions challenges the common practice of pooling all omega-6 PUFAs into a single, global omega-6 biomarker. Not all omega-6 FAs exert similar metabolic effects [49]. Hence, investigating individual omega-6 FAs, rather than treating omega-6 PUFAs as a homogeneous group should be prioritized. Future investigations to elucidate how non-LA omega-6 PUFAs may influence adiposity, including their roles in eicosanoid production and inflammatory signaling, are warranted.

Regardless, our findings support current dietary guidelines [50] that emphasize LA-rich foods, such as nuts, seeds, soybean, corn, and sunflower oils, as part of a balanced diet for weight maintenance.

The non-linear associations between LA and adiposity are interesting: cross-sectional analyses suggest threshold effects, with the strongest inverse associations observed up to ~35% of total fatty acids, beyond which additional LA confers little incremental benefit. This pattern is consistent with the behaviors of other FAs [51], where endogenous regulation limits saturation and excess accumulation [7]. In contrast, longitudinal analysis reveals a non-linear curve, suggesting that higher LA levels beyond this threshold may still confer benefits for changes in weight and whole-body fat mass, though replication in other studies are needed to confirm this finding. Interactions by age and sex indicate heterogeneity in effect sizes, underscoring the need to consider demographic stratification in future investigations.

Several studies have examined the association between omega-6 PUFAs and body anthropometrics, yet findings remain inconsistent. In analyses based on dietary intake, total omega-6 PUFAs, LA, and AA were generally not associated with indices of adiposity, weight gain, body fat, and relative fat mass [28, 31–34]. An exception is a cross-sectional US national survey, which reported an inverse association between total omega-6 intake and body fat percentage [52]. Studies using circulating biomarkers provide a somewhat clearer picture. One study which examined levels of cholesterol ester LA found significant inverse associations with sagittal abdominal diameter, waist circumference, and waist–hip ratio [25], supporting our main findings. By contrast, higher levels of circulating LA was positively linked to lean tissue volume [18, 23] and skeletal muscle mass [24] which diverges from our secondary findings on fat-free mass. Though, notably we estimated fat-free mass via bioimpedance rather than dual-energy X-ray absorptiometry (DXA). Only one study evaluated both cross-sectional and longitudinal associations [26]. LA, but not total omega-6 PUFAs or AA, was inversely associated with BMI cross-sectionally, whereas longitudinally (with a linear assumption), total omega-6 PUFAs was positively associated with BMI change, with null findings for LA and AA [26]. Such inconsistencies across studies likely reflect variation in exposure assessment (dietary vs. circulating biomarkers), outcome measurement (DXA vs. bioimpedance) and population characteristics. Against this backdrop, our study – the largest to date to our knowledge – extends existing evidence by examining both cross-sectional and longitudinal associations of omega-6 PUFAs, particularly LA, with seven anthropometric outcomes, thereby addressing a key knowledge gap.

Several strengths warrant emphasis. First, our investigation benefited from a substantial sample size (over a quarter million cross-sectionally), which provided robust statistical power to detect associations. Second, the use of objective fatty acid biomarkers rather than self-reported intake minimized reporting bias, strengthening the validity of exposure assessment. Third, comprehensively characterized exposures, outcomes, and potential confounding variables measured via standardized protocols enhanced comparability and reproducibility. Lastly, the availability of repeated anthropometric measurements allowed us to assess associations in both cross-sectional and longitudinal frameworks, providing insights into both baseline relationships and prospective changes over time.

Nonetheless, some limitations should be acknowledged. Non-LA omega-6 FAs were not individually quantified, precluding a more detailed investigation of their specific roles. Bioimpedance is less precise compared to DXA, but these methos have been shown to be strongly correlated at a population level [53]. Participants were predominantly middle-aged to older adults, possibly limiting generalizability to younger populations. The UKBB also predominantly consists of White individuals and those with a higher socioeconomic status; while these factors were accounted for in the analyses, some degree of selection bias may remain. As with all observational research, causality cannot be inferred, and residual confounding cannot be entirely excluded. However, our findings may provide insights to inform future randomized controlled trials and interventions studies.

In conclusion, higher levels of total omega-6 PUFAs, particularly LA, were associated with smaller waist circumference and lower weight and whole-body fat mass in both cross-sectional and longitudinal analyses. In contrast, non-LA omega-6 demonstrated associations in the opposite direction with weak effect sizes. Our findings support dietary recommendations to include LA-rich oils in the diet, rather than discouraging their intake. Furthermore, the divergent patterns between LA and non-LA omega-6 highlight the limitations of relying on a global total omega-6 biomarker, underscoring the need for future studies to investigate the specific determinants and potential health implications of individual non-LA omega-6 fatty acids.

## Abbreviations

AA: arachidonic acid
BMI: body mass index
DiHOME: dihydroxy-9Z-octadecenoic acid
DLGA: dihomo-γ-linolenic acid
DXA: dual-energy X-ray absorptiometry
EpOME: epoxyoctadecenoic acid
GLA: γ-linolenic acid
IQ_5_R: Interquintile range
LA: linoleic acid
PPAR: peroxisome proliferator–activated receptors
PUFA: polyunsaturated fatty acid(s)
Q: Quintile
UKBB: UK Biobank

## Author contributions

The authors’ responsibilities were as follows – WSH and NLT: conceived the project; NLT: gained access to the UK Biobank dataset; JW, EDJ, and NLT: analyzed the data; HTML: prepared the first draft, which JW, NLT, MAB, and WSH improved by critical review; HTML and WSH: had primary responsibility for final content; and all authors: read and approved the final manuscript.

## Data availability

Data described in the manuscript, code book, and analytic code will be made available upon request pending application and approval.

