## Supplemental Tables and Figures for "Omega-6 polyunsaturated fatty acids and adiposity in the UK Biobank Cohort: a cross-sectional and longitudinal prospective analysis"

**Supplementary Table 1.** Definition of covariates in the UK Biobank

| Variable | UK Biobank variable name | Definition/categories |
| --- | --- | --- |
| Age | 31003 | Continuous in years |
| Sex | 31 | Categories:<br>Female<br>Male |
| Level of education | 6138 | Categories:<br>Less than high school<br>High school<br>College<br>Missing |
| Townsend Deprivation Index | 22189 | Continuous |
| Race | 21000 | Categories:<br>White<br>Black<br>Asian<br>Missing<br>Other |
| Physical Activity | 874, 894, 914, 864, 884, 904 | Categories:<br>Lowest<br>Low<br>High<br>Highest<br>Missing |
| Weight | 21002 | Continuous variable in kg |
| Standing Height | 50 | Continuous variable in cm |
| Waist Circumference | 48 | Continuous variable in cm |
| Whole Body Fat Mass | 23100 | Continuous variable in kg |
| Whole Body Fat Free Mass | 23101 | Continuous variable in kg |
| Trunk Fat Mass | 23128 | Continuous variable in kg |
| Trunk Percent Fat | 23127 | Continuous variable in percent |
| Linoleic Acid to total fatty acids percentage | 23456 | Continuous variable in percent of total fatty acids |
| Docosahexaenoic acid to total fatty acids percentage | 23457 | Continuous variable in percent of total fatty acids |
| Non-linoleic omega-6 fatty acids | Obtained by subtracting Linoleic acid (variable 23456) from Omega-6 Fatty Acids to Total Fatty Acids percentage (variable 23452) | Continuous variable in percent of total fatty acids |
| Omega-6 fatty acids to total fatty acids percentage | 23452 | Continuous variable in percent of total fatty acids |

**Supplementary Table 2. Partial correlations between seven anthropometric measurements in the UK Biobank**

| **Cross-sectional correlations** | WC | Weight | BMI | WB FFM | WB FM | Trunk FM | Trunk % fat |
|---:|---|---|---|---|---|---|---|
| WC | 1.00 | 0.89 | 0.81 | 0.68 | 0.65 | 0.77 | 0.43 |
| Weight | 0.89 | 1.00 | 0.83 | 0.80 | 0.69 | 0.81 | 0.40 |
| BMI | 0.81 | 0.83 | 1.00 | 0.41 | 0.88 | 0.87 | 0.66 |
| WB FFM | 0.68 | 0.80 | 0.41 | 1.00 | 0.12 | 0.33 | -0.19 |
| WB FM | 0.65 | 0.69 | 0.88 | 0.12 | 1.00 | 0.94 | 0.88 |
| Trunk FM | 0.77 | 0.81 | 0.87 | 0.33 | 0.94 | 1.00 | 0.84 |
| Trunk % fat | 0.43 | 0.40 | 0.66 | -0.19 | 0.88 | 0.84 | 1.00 |
| **Longitudinal correlations** | WC | Weight | BMI | WB FFM | WB FM | Trunk FM | Trunk % fat |
| WC | 1.00 | 0.61 | 0.61 | 0.34 | 0.55 | 0.50 | 0.41 |
| Weight | 0.61 | 1.00 | 0.97 | 0.62 | 0.86 | 0.75 | 0.57 |
| BMI | 0.61 | 0.97 | 1.00 | 0.56 | 0.86 | 0.73 | 0.57 |
| WB FFM | 0.34 | 0.62 | 0.56 | 1.00 | 0.25 | 0.11 | -0.14 |
| WB FM | 0.55 | 0.86 | 0.86 | 0.25 | 1.00 | 0.94 | 0.84 |
| Trunk FM | 0.50 | 0.75 | 0.73 | 0.11 | 0.94 | 1.00 | 0.93 |
| Trunk % fat | 0.41 | 0.57 | 0.57 | -0.14 | 0.84 | 0.93 | 1.00 |

Abbreviations: Body mass index, BMI; Fat mass, FM; Fat-free mass, FFM; Whole-body, WB; Waist circumference, WC

**Supplementary Table 3. The cross-sectional association between linoleic acid, non-linoleic acid omega-6, and total omega-6 polyunsaturated fatty acid and waist circumference, weight, and whole-body fat mass in the UK Biobank**

| | Quintiles of fatty acid | | | | | | | |
|---|---|---|---|---|---|---|---|---|
| *Linoleic acid* | I | II | III | IV | V | p- trend[1] | Per IQ5R[2] | p-value |
| Total N | 54,516 | 54,483 | 54,537 | 54,530 | 54,521 | | 272,587 | |
| Median (% total fatty acid) | 24.4 | 27.3 | 29.1 | 30.8 | 33.0 | | 29.1 | |
| Waist circumference (cm) [3] | 0 (Ref) | -3.13 (-3.26, -3.00) | -5.75 (-5.88, -5.62) | -8.10 (-8.23, -7.97) | -11.04 (-11.17, -10.91) | <0.001 | -9.75 (-9.86, -9.65) | <0.001 |
| Total N | 54,516 | 54,483 | 54,537 | 54,530 | 54,521 | | 272,587 | |
| Median (% total fatty acid) | 24.4 | 27.3 | 29.1 | 30.8 | 33.0 | | 29.1 | |
| Weight (kg) [3] | 0 (Ref) | -3.20 (-3.34, -3.05) | -5.89 (-6.04, -5.75) | -8.39 (-8.53, -8.24) | -11.77 (-11.92, -11.62) | <0.001 | -10.38 (-10.50, -10.26) | <0.001 |
| Total N | 53,395 | 53,544 | 53,692 | 53,732 | 53,647 | | 268,010 | |
| Median (% total fatty acid) | 24.4 | 27.3 | 29.1 | 30.8 | 33.0 | | 29.1 | |
| Whole-body fat mass (kg) [3] | 0 (Ref) | -1.95 (-2.05, -1.85) | -3.77 (-3.87, -3.67) | -5.51 (-5.61, -5.41) | -7.87 (-7.97, -7.77) | <0.001 | -6.93 (-7.02, -6.85) | <0.001 |
| *Non-LA omega-6* | | | | | | | | |
| Total N | 54,515 | 54,507 | 54,520 | 54,520 | 54,525 | | 272,587 | |
| Median (% total fatty acid) | 6.6 | 7.9 | 8.8 | 9.8 | 11.4 | | 8.8 | |
| Waist circumference (cm) [3] | 0 (Ref) | 0.08 (-0.05, 0.21) | 0.10 (-0.04, 0.24) | 0.24 (0.10, 0.38) | 1.46 (1.32, 1.61) | <0.001 | 1.55 (1.43, 1.67) | <0.001 |
| Total N | 54,515 | 54,507 | 54,520 | 54,520 | 54,525 | | 272,587 | |
| Median (% total fatty acid) | 6.6 | 7.9 | 8.8 | 9.8 | 11.4 | | 8.8 | |
| Weight (kg) [3] | 0 (Ref) | 0.47 (0.32, 0.62) | 0.57 (0.41, 0.72) | 0.95 (0.79, 1.10) | 2.41 (2.25, 2.58) | <0.001 | 2.41 (2.28, 2.55) | <0.001 |
| Total N | 53,723 | 53,718 | 53,639 | 53,590 | 53,340 | | 268,010 | |
| Median (% total fatty acid) | 6.6 | 7.9 | 8.8 | 9.8 | 11.4 | | 8.8 | |
| Whole-body fat mass (kg) [3] | 0 (Ref) | 0.38 (0.27, 0.48) | 0.51 (0.40, 0.61) | 0.77 (0.66, 0.88) | 1.81 (1.69, 1.92) | <0.001 | 1.78 (1.69, 1.88) | <0.001 |
| *Total omega-6* | | | | | | | | |
| Total N | 54,518 | 54,505 | 54,500 | 54,534 | 54,530 | | 272,587 | |
| Median (% total fatty acid) | 32.9 | 36.4 | 38.4 | 40.0 | 42.0 | | 38.4 | |
| Waist circumference (cm) [3] | 0 (Ref) | -2.62 (-2.75, -2.49) | -5.10 (-5.23, -4.97) | -7.64 (-7.77, -7.51) | -10.24 (-10.37, -10.10) | <0.001 | -9.00 (-9.11, -8.90) | <0.001 |
| Total N | 54,518 | 54,505 | 54,500 | 54,534 | 54,530 | | 272,587 | |
| Median (% total fatty acid) | 32.9 | 36.4 | 38.4 | 40.0 | 42.0 | | 38.4 | |

| | | | | | | | | |
|---|---|---|---|---|---|---|---|---|
| Weight (kg) [3] | 0 (Ref) | -2.66 (-2.81, -2.51) | -5.08 (-5.23, -4.93) | -7.66 (-7.81, -7.51) | -10.51 (-10.66, -10.36) | <0.001 | -9.22 (-9.34, -9.09) | <0.001 |
| Total N | 53,532 | 53,593 | 53,695 | 53,655 | 53,535 | | 268,010 | |
| Median (% total fatty acid) | 32.9 | 36.4 | 38.4 | 40.0 | 42.0 | | 38.4 | |
| Whole-body fat mass (kg) [3] | 0 (Ref) | -1.54 (-1.65, -1.44) | -3.19 (-3.29, -3.08) | -4.97 (-5.07, -4.86) | -6.97 (-7.07, -6.86) | <0.001 | -6.07 (-6.16, -5.98) | <0.001 |

Models are adjusted for age, sex, race, standing height, education, Townsend scores, physical activity, and levels of DHA.

[1] P-trend is generated by assigning participants the median value in each quintile then assessing quintiles as continuous variables

[2] Interquintile range (IQ5R) is the difference between the first and fifth quintiles

[3] Findings are reported as the difference in comparison to the lowest quintile. Negative values mean that the outcome of interest is smaller compared to the reference group (set as zero), while positive values indicate the opposite.

**Supplementary Table 4: The longitudinal association between total omega-6 polyunsaturated fatty acid and waist circumference, weight, and whole-body fat mass in the UK Biobank**

| | Quintiles of fatty acid | | | | | | | |
|---|---|---|---|---|---|---|---|---|
| *Total omega-6* | I | II | III | IV | V | p- trend[1] | Per IQ5R[2] | p-value |
| Total N | 11,658 | 11,680 | 11,685 | 11,668 | 11,644 | | 58,335 | |
| Median (% total fatty acid) | 33.4 | 36.8 | 38.7 | 40.3 | 42.2 | | 38.7 | |
| Waist circumference (cm) [3] | 0 (Ref) | -0.01 (-0.03, 0.02) | -0.02 (-0.04, 0.00) | -0.05 (-0.07, -0.03) | -0.06 (-0.09, -0.04) | <0.001 | -0.05 (-0.07, -0.03) | <0.001 |
| Total N | 11,658 | 11,680 | 11,685 | 11,668 | 11,644 | | 58,335 | |
| Median (% total fatty acid) | 33.4 | 36.8 | 38.7 | 40.3 | 42.2 | | 38.7 | |
| Weight (kg) [3] | 0 (Ref) | 0.02 (0.01, 0.04) | 0.04 (0.02, 0.05) | 0.03 (0.01, 0.05) | 0.02 (0.00, 0.04) | 0.075 | 0.03 (0.01, 0.05) | <0.001 |
| Total N | 8,756 | 8,769 | 8,750 | 8,472 | 8,358 | | 43,105 | |
| Median (% total fatty acid) | 33.4 | 36.8 | 38.7 | 40.3 | 42.2 | | 38.7 | |
| Whole-body fat mass (kg) [3] | 0 (Ref) | 0.02 (0.00, 0.04) | 0.02 (0.00, 0.04) | 0.03 (0.01, 0.05) | 0.02 (0.00, 0.04) | 0.046 | 0.03 (0.01, 0.04) | 0.001 |

Models are adjusted for age, sex, race, standing height, baseline weight, baseline waist circumference, baseline whole body fat mass, education, Townsend scores, physical activity, and levels of DHA.

[1] P-trend is generated by assigning participants the median value in each quintile then assessing quintiles as continuous variables

[2] Interquintile range (IQ5R) is the difference between the first and fifth quintiles

[3] Findings are reported as the difference in comparison to the lowest quintile in terms of change per year. Negative values mean that the outcome of interest is smaller compared to the reference group (set as zero), while positive values indicate the opposite.

**Supplementary Table 5: The cross-sectional association between linoleic acid, non-linoleic acid omega-6, and total omega-6 polyunsaturated fatty acid and body mass index, whole-body fat-free mass, trunk fat mass, and trunk percent fat in the UK Biobank**

| | Quintiles of fatty acid | | | | | | | |
|---|---|---|---|---|---|---|---|---|
| *Linoleic acid* | I | II | III | IV | V | p- trend [1] | Per IQ5R [2] | p-value |
| Total N | 54,516 | 54,483 | 54,537 | 54,530 | 54,521 | | 272,587 | |
| Median (% total fatty acid) | 24.4 | 27.3 | 29.1 | 30.8 | 33.0 | | 29.1 | |
| Body mass index (kg/m$^2$) [3] | 0 (Ref) | -1.09 (-1.15, -1.04) | -2.04 (-2.09, -1.99) | -2.92 (-2.98, -2.87) | -4.12 (-4.17, -4.07) | <0.001 | -3.63 (-3.67, -3.59) | <0.001 |
| Total N | 53,403 | 53,578 | 53,758 | 53,837 | 53,848 | | 268,424 | |
| Median (% total fatty acid) | 24.4 | 27.3 | 29.1 | 30.8 | 33.0 | | 29.1 | |
| Whole-body fat-free mass (kg) [3] | 0 (Ref) | -1.21 (-1.27, -1.15) | -2.07 (-2.13, -2.01) | -2.81 (-2.87, -2.75) | -3.79 (-3.86, -3.73) | <0.001 | -3.38 (-3.43, -3.33) | <0.001 |
| Total N | 53,378 | 53,544 | 53,723 | 53,806 | 53,802 | | 268,253 | |
| Median (% total fatty acid) | 24.4 | 27.3 | 29.1 | 30.8 | 33.0 | | 29.1 | |
| Trunk fat mass (kg) [3] | 0 (Ref) | -1.06 (-1.11, -1.00) | -2.05 (-2.10, -1.99) | -3.00 (-3.06, -2.94) | -4.32 (-4.38, -4.27) | <0.001 | -3.81 (-3.86, -3.77) | <0.001 |
| Total N | 53,379 | 53,549 | 53,726 | 53,807 | 53,807 | | 268,268 | |
| Median (% total fatty acid) | 24.4 | 27.3 | 29.1 | 30.8 | 33.0 | | 29.1 | |
| Trunk percent fat (%) [3] | 0 (Ref) | -1.16 (-1.24, -1.08) | -2.40 (-2.48, -2.32) | -3.70 (-3.78, -3.62) | -5.65 (-5.74, -5.57) | <0.001 | -4.96 (-5.02, -4.89) | <0.001 |
| *Non-LA omega-6* | | | | | | | | |
| Total N | 54,515 | 54,507 | 54,520 | 54,520 | 54,525 | | 272,587 | |
| Median (% total fatty acid) | 6.6 | 7.9 | 8.8 | 9.8 | 11.4 | | 8.8 | |
| Body mass index (kg/m$^2$) [3] | 0 (Ref) | 0.17 (0.12, 0.23) | 0.22 (0.17, 0.28) | 0.36 (0.30, 0.42) | 0.89 (0.83, 0.95) | <0.001 | 0.89 (0.84, 0.93) | <0.001 |
| Total N | 53,779 | 53,777 | 53,725 | 53,694 | 53,449 | | 268,424 | |
| Median (% total fatty acid) | 6.6 | 7.9 | 8.8 | 9.8 | 11.4 | | 8.8 | |
| Whole-body fat-free mass (kg) [3] | 0 (Ref) | 0.11 (0.05, 0.17) | 0.07 (0.01, 0.13) | 0.18 (0.12, 0.25) | 0.59 (0.52, 0.65) | <0.001 | 0.60 (0.55, 0.65) | <0.001 |
| Total N | 53,754 | 53,736 | 53,693 | 53,661 | 53,409 | | 268,253 | |
| Median (% total fatty acid) | 6.6 | 7.9 | 8.8 | 9.8 | 11.4 | | 8.8 | |
| Trunk fat mass (kg) [3] | 0 (Ref) | 0.20 (0.14, 0.26) | 0.26 (0.20, 0.32) | 0.40 (0.34, 0.46) | 0.97 (0.90, 1.03) | <0.001 | 0.96 (0.90, 1.01) | <0.001 |
| Total N | 53,754 | 53,739 | 53,697 | 53,664 | 53,414 | | 268,268 | |
| Median (% total fatty acid) | 6.6 | 7.9 | 8.8 | 9.8 | 11.4 | | 8.8 | |
| Trunk percent fat (%) [3] | 0 (Ref) | 0.19 (0.10, 0.27) | 0.24 (0.15, 0.32) | 0.38 (0.30, 0.47) | 1.10 (1.01, 1.19) | <0.001 | 1.10 (1.03, 1.17) | <0.001 |

| | | | | | | | | |
|---|---|---|---|---|---|---|---|---|
| *Total omega-6* | | | | | | | | |
| Total N | 54,518 | 54,505 | 54,500 | 54,534 | 54,530 | | 272,587 | |
| Median (% total fatty acid) | 32.9 | 36.4 | 38.4 | 40.0 | 42.0 | | 38.4 | |
| Body mass index (kg/$m^2$) [3] | 0 (Ref) | -0.90 (-0.95, -0.85) | -1.76 (-1.81, -1.71) | -2.67 (-2.72, -2.61) | -3.66 (-3.71, -3.60) | <0.001 | -3.20 (-3.25, -3.16) | <0.001 |
| Total N | 53,538 | 53,610 | 53,755 | 53,758 | 53,763 | | 268,424 | |
| Median (% total fatty acid) | 32.9 | 36.4 | 38.4 | 40.0 | 42.0 | | 38.4 | |
| Whole-body fat-free mass (kg) [3] | 0 (Ref) | -1.11 (-1.17, -1.04) | -1.87 (-1.93, -1.81) | -2.66 (-2.72, -2.60) | -3.46 (-3.52, -3.40) | <0.001 | -3.09 (-3.14, -3.04) | <0.001 |
| Total N | 53,515 | 53,585 | 53,720 | 53,719 | 53,714 | | 268,253 | |
| Median (% total fatty acid) | 32.9 | 36.4 | 38.4 | 40.0 | 42.0 | | 38.4 | |
| Trunk fat mass (kg) [3] | 0 (Ref) | -0.83 (-0.89, -0.77) | -1.72 (-1.78, -1.66) | -2.70 (-2.76, -2.64) | -3.85 (-3.90, -3.79) | <0.001 | -3.35 (-3.40, -3.30) | <0.001 |
| Total N | 53,516 | 53,589 | 53,722 | 53,721 | 53,720 | | 268,268 | |
| Median (% total fatty acid) | 32.9 | 36.4 | 38.4 | 40.0 | 42.0 | | 38.4 | |
| Trunk percent fat (%) [3] | 0 (Ref) | -0.88 (-0.96, -0.79) | -2.03 (-2.11, -1.95) | -3.40 (-3.49, -3.32) | -5.14 (-5.23, -5.06) | <0.001 | -4.43 (-4.49, -4.36) | <0.001 |

Models are adjusted for age, sex, race, standing height, education, Townsend scores, physical activity, and levels of DHA.

[1] P-trend is generated by assigning participants the median value in each quintile then assessing quintiles as continuous variables

[2] Interquintile range (IQ5R) is the difference between the first and fifth quintiles

[3] Findings are reported as the difference in comparison to the lowest quintile. Negative values mean that the outcome of interest is smaller compared to the reference group (set as zero), while positive values indicate the opposite.

**Supplementary Table 6: The longitudinal association between linoleic acid, non-linoleic acid omega-6, and total omega-6 polyunsaturated fatty acid and change in body mass index, whole-body fat-free mass, trunk fat mass, and trunk percent fat in the UK Biobank**

| | Quintiles of fatty acid | | | | | | | |
|---|---|---|---|---|---|---|---|---|
| *Linoleic acid* | I | II | III | IV | V | p- trend [1] | Per IQ5R [2] | p-value |
| Total N | 11,654 | 11,657 | 11,689 | 11,679 | 11,656 | | 58,335 | |
| Median (% total fatty acid) | 25.0 | 27.8 | 29.5 | 31.1 | 33.3 | | 29.5 | |
| Body mass index ($kg/m^2$) [3] | 0 (Ref) | 0.01 (0.00, 0.01) | 0.00 (0.00, 0.01) | 0.00 (-0.01, 0.01) | -0.01 (-0.02, -0.01) | <0.001 | -0.01 (-0.02, -0.01) | <0.001 |
| Total N | 8,873 | 8,807 | 8,616 | 8,506 | 8,319 | | 43,121 | |
| Median (% total fatty acid) | 25.0 | 27.7 | 29.5 | 31.1 | 33.3 | | 29.4 | |
| Whole-body fat-free mass (kg) [3] | 0 (Ref) | 0.01 (0.00, 0.02) | 0.01 (-0.01, 0.02) | 0.01 (0.00, 0.02) | 0.00 (-0.01, 0.01) | 0.400 | 0.00 (-0.01, 0.01) | 0.448 |
| Total N | 8,868 | 8,801 | 8,613 | 8,496 | 8,308 | | 43,086 | |
| Median (% total fatty acid) | 25.0 | 27.7 | 29.5 | 31.1 | 33.3 | | 29.4 | |
| Trunk fat mass (kg) [3] | 0 (Ref) | 0.01 (0.00, 0.02) | 0.00 (-0.01, 0.02) | 0.00 (-0.02, 0.01) | -0.03 (-0.04, -0.01) | <0.001 | -0.02 (-0.03, -0.01) | <0.001 |
| Total N | 8,868 | 8,802 | 8,614 | 8,496 | 8,310 | | 43,090 | |
| Median (% total fatty acid) | 25.0 | 27.7 | 29.5 | 31.1 | 33.3 | | 29.4 | |
| Trunk percent fat (%) [3] | 0 (Ref) | 0.01 (-0.01, 0.03) | 0.00 (-0.02, 0.02) | -0.02 (-0.03, 0.00) | -0.04 (-0.06, -0.02) | <0.001 | -0.04 (-0.05, -0.02) | <0.001 |
| *Non-LA omega-6* | | | | | | | | |
| Total N | 11,668 | 11,692 | 11,667 | 11,665 | 11,643 | | 58,335 | |
| Median (% total fatty acid) | 6.6 | 7.9 | 8.8 | 9.8 | 11.3 | | 8.8 | |
| Body mass index ($kg/m^2$) [3] | 0 (Ref) | 0.01 (0.01, 0.02) | 0.02 (0.01, 0.03) | 0.02 (0.02, 0.03) | 0.05 (0.04, 0.05) | <0.001 | 0.04 (0.04, 0.05) | <0.001 |
| Total N | 8,547 | 8,606 | 8,539 | 8,619 | 8,810 | | 43,121 | |
| Median (% total fatty acid) | 6.7 | 7.9 | 8.8 | 9.8 | 11.3 | | 8.8 | |
| Whole-body fat-free mass (kg) [3] | 0 (Ref) | 0.00 (-0.01, 0.01) | 0.00 (-0.01, 0.01) | 0.00 (-0.01, 0.01) | 0.02 (0.01, 0.04) | <0.001 | 0.02 (0.01, 0.03) | <0.001 |
| Total N | 8,543 | 8,596 | 8,533 | 8,612 | 8,802 | | 43,086 | |
| Median (% total fatty acid) | 6.7 | 7.9 | 8.8 | 9.8 | 11.3 | | 8.8 | |
| Trunk fat mass (kg) [3] | 0 (Ref) | 0.02 (0.01, 0.03) | 0.04 (0.02, 0.05) | 0.04 (0.03, 0.05) | 0.07 (0.06, 0.08) | <0.001 | 0.07 (0.06, 0.08) | <0.001 |
| Total N | 8,543 | 8,596 | 8,534 | 8,612 | 8,805 | | 43,090 | |
| Median (% total fatty acid) | 6.7 | 7.9 | 8.8 | 9.8 | 11.3 | | 8.8 | |
| Trunk percent fat (%) [3] | 0 (Ref) | 0.03 (0.01, 0.05) | 0.06 (0.04, 0.08) | 0.07 (0.05, 0.09) | 0.11 (0.09, 0.13) | <0.001 | 0.11 (0.09, 0.12) | <0.001 |

| | | | | | | | | |
|---|---|---|---|---|---|---|---|---|
| *Total omega-6* | | | | | | | | |
| Total N | 11,658 | 11,680 | 11,685 | 11,668 | 11,644 | | 58,335 | |
| Median (% total fatty acid) | 33.4 | 36.8 | 38.7 | 40.3 | 42.2 | | 38.7 | |
| Body mass index ($kg/m^2$) [3] | 0 (Ref) | 0.01 (0.00, 0.01) | 0.01 (0.01, 0.02) | 0.01 (0.01, 0.02) | 0.01 (0.00, 0.02) | 0.004 | 0.01 (0.01, 0.02) | <0.001 |
| Total N | 8,757 | 8,769 | 8,750 | 8,476 | 8,369 | | 43,121 | |
| Median (% total fatty acid) | 33.4 | 36.8 | 38.7 | 40.3 | 42.2 | | 38.7 | |
| Whole-body fat-free mass (kg) [3] | 0 (Ref) | 0.01 (0.00, 0.02) | 0.02 (0.01, 0.03) | 0.01 (0.00, 0.02) | 0.01 (0.00, 0.03) | 0.040 | 0.02 (0.01, 0.02) | 0.001 |
| Total N | 8,751 | 8,767 | 8,742 | 8,466 | 8,360 | | 43,086 | |
| Median (% total fatty acid) | 33.4 | 36.8 | 38.7 | 40.3 | 42.2 | | 38.7 | |
| Trunk fat mass (kg) [3] | 0 (Ref) | 0.01 (0.00, 0.02) | 0.01 (0.00, 0.03) | 0.02 (0.00, 0.03) | 0.01 (0.00, 0.03) | 0.034 | 0.02 (0.01, 0.03) | 0.001 |
| Total N | 8,751 | 8,767 | 8,742 | 8,468 | 8,362 | | 43,090 | |
| Median (% total fatty acid) | 33.4 | 36.8 | 38.7 | 40.3 | 42.2 | | 38.7 | |
| Trunk percent fat (%) [3] | 0 (Ref) | 0.01 (-0.01, 0.03) | 0.01 (-0.01, 0.03) | 0.02 (0.00, 0.04) | 0.02 (0.00, 0.04) | 0.085 | 0.02 (0.01, 0.04) | 0.008 |

Models are adjusted for age, sex, race, standing height, baseline weight, baseline waist circumference, baseline whole body fat mass, education, Townsend scores, physical activity, and levels of DHA.

[1] P-trend is generated by assigning participants the median value in each quintile then assessing quintiles as continuous variables

[2] Interquintile range (IQ5R) is the difference between the first and fifth quintiles

[3] Findings are reported as the difference in comparison to the lowest quintile in terms of change per year. Negative values mean that the outcome of interest is smaller compared to the reference group (set as zero), while positive values indicate the opposite.

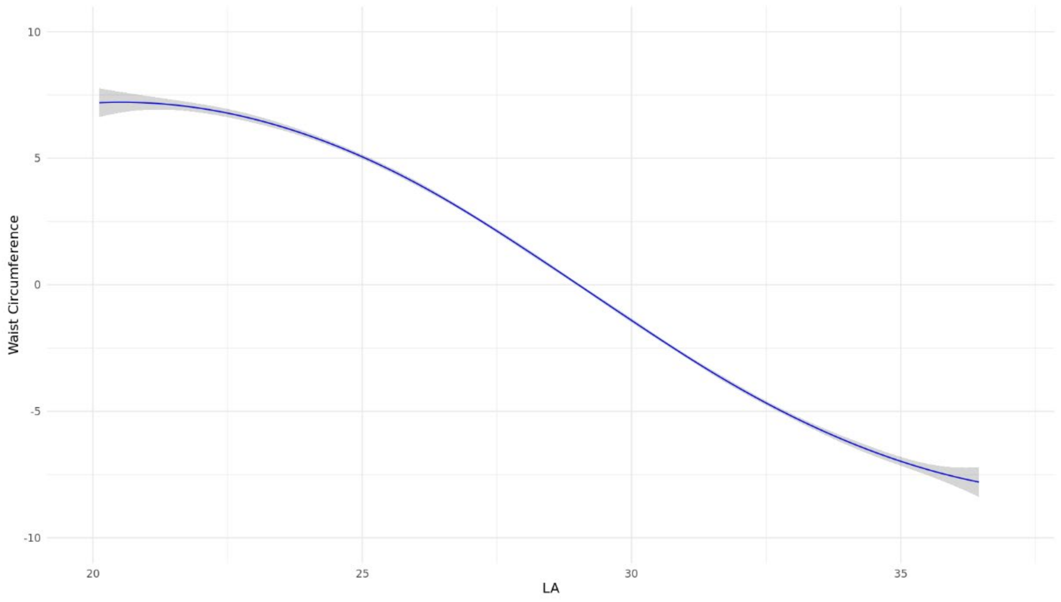


**Supplementary Figure 1: Restricted cubic splines demonstrating the non-linear cross-sectional association between linoleic acid (LA) and waist circumference (cm).** The blue curve represents the fitted spline effect, and the shaded area indicates the 95% confidence interval (CI).

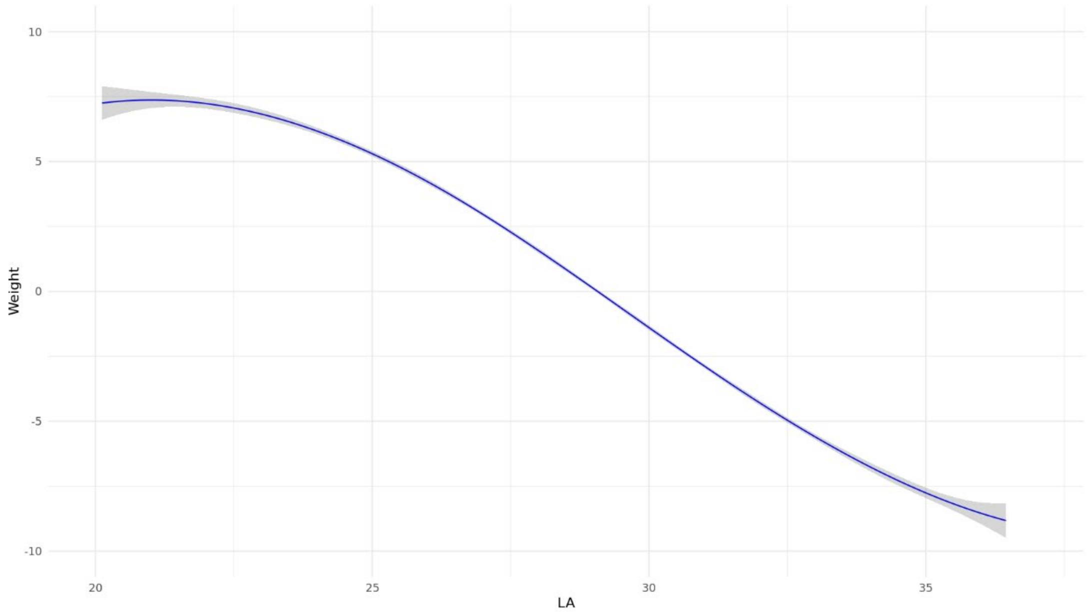


**Supplementary Figure 2: Restricted cubic splines demonstrating the non-linear cross-sectional association between linoleic acid (LA) and weight (kg).** The blue curve represents the fitted spline effect, and the shaded area indicates the 95% confidence interval (CI).

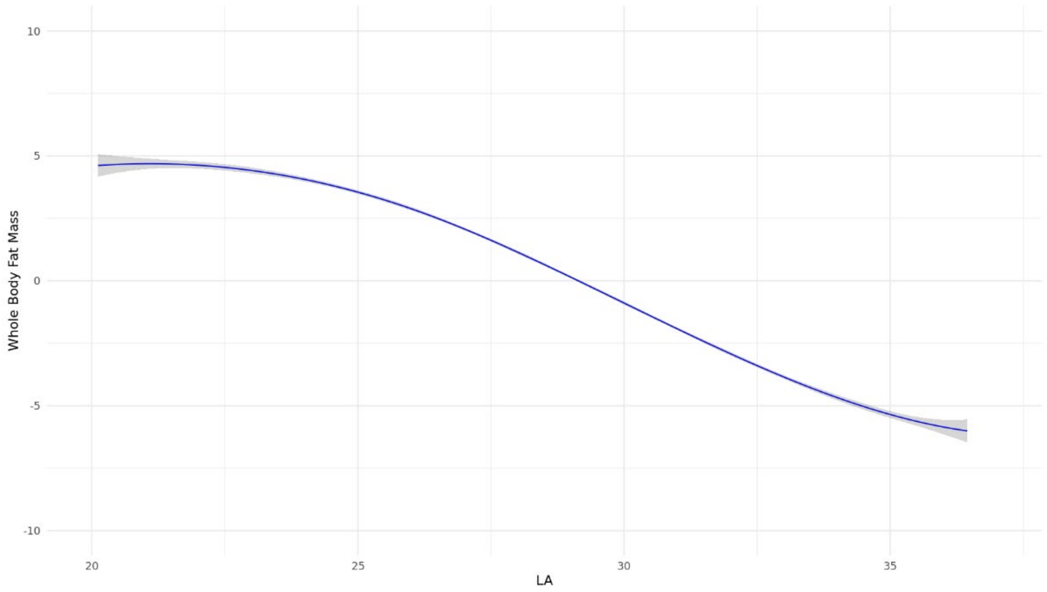


**Supplementary Figure 3: Restricted cubic splines demonstrating the non-linear cross-sectional association between linoleic acid (LA) and whole-body fat mass (kg).** The blue curve represents the fitted spline effect, and the shaded area indicates the 95% confidence interval (CI).

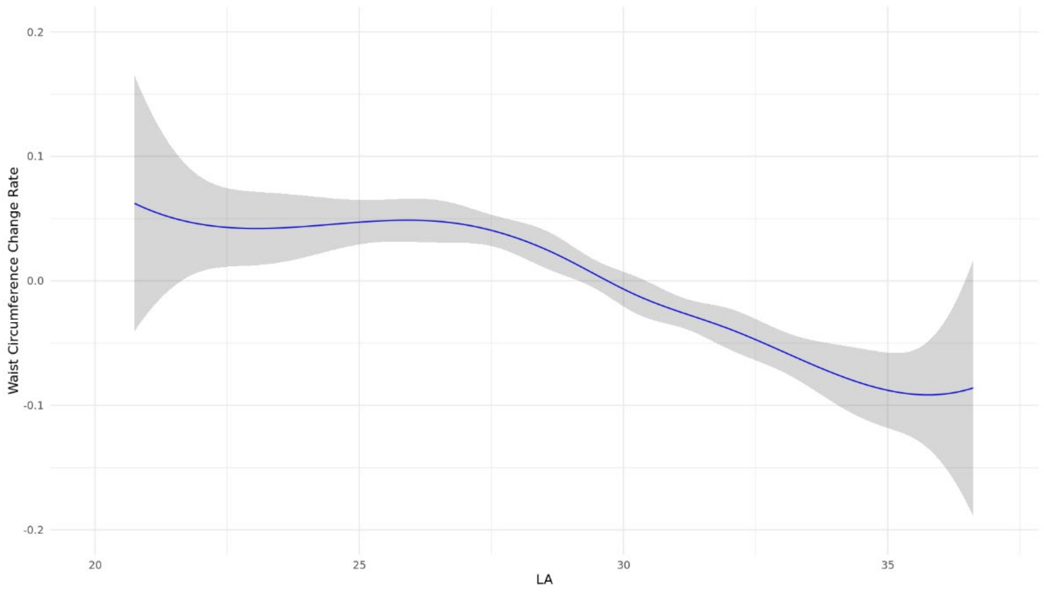


**Supplementary Figure 4: Restricted cubic splines demonstrating the non-linear longitudinal association between linoleic acid (LA) and waist circumference (change in cm per year).** The blue curve represents the fitted spline effect, and the shaded area indicates the 95% confidence interval (CI).

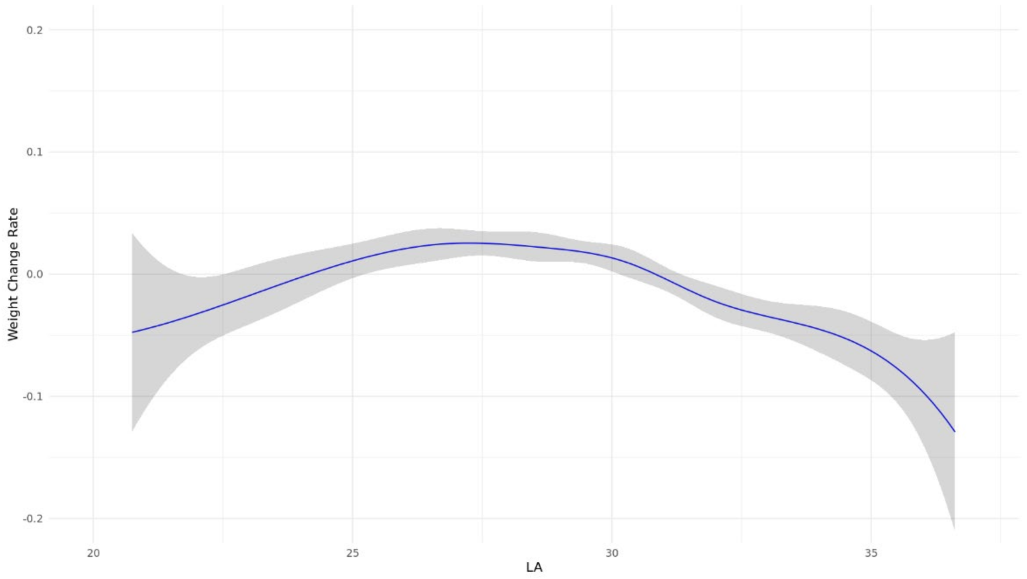


**Supplementary Figure 5: Restricted cubic splines demonstrating the non-linear longitudinal association between linoleic acid (LA) and weight (change in kg per year).** The blue curve represents the fitted spline effect, and the shaded area indicates the 95% confidence interval (CI).

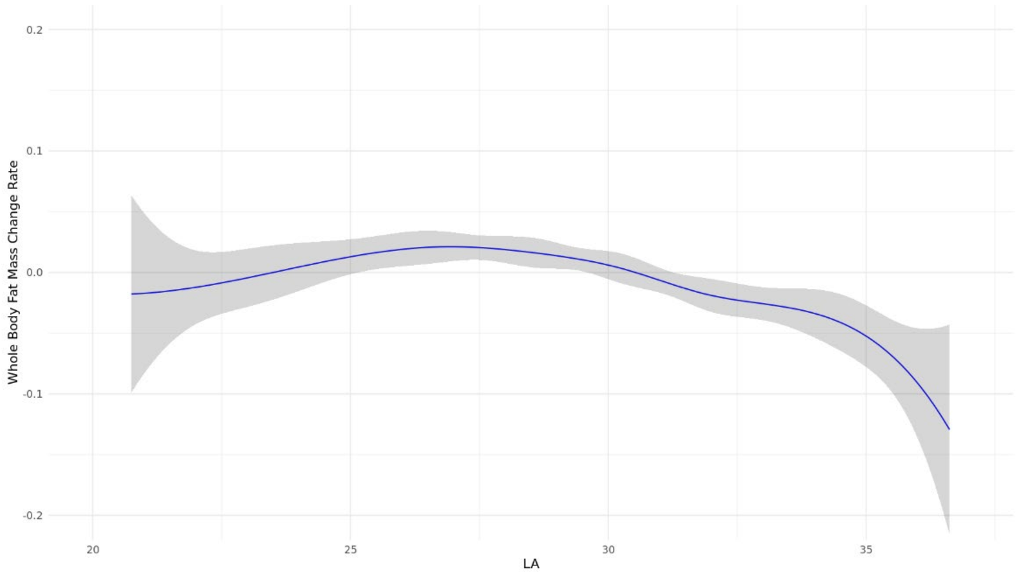


**Supplementary Figure 6: Restricted cubic splines demonstrating the non-linear longitudinal association between linoleic acid (LA) and whole body fat mass (change in kg per year).** The blue curve represents the fitted spline effect, and the shaded area indicates the 95% confidence interval (CI).

**Supplementary Table 7: Cross-sectional analyses between linoleic acid (% total fatty acid) expressed as per inter-quintile range and waist circumference, weight, and whole-body fat mass stratified by age and sex**

| | Waist circumference (cm) [1] | p-value [2] | Weight (kg) [1] | p-value [2] | Whole-body fat mass (kg) [1] | p-value [2] |
|---|---|---|---|---|---|---|
| *Age* | | | | | | |
| >60 years | -9.50 (-9.66, -9.35) | | -9.67 (-9.84, -9.51) | | -6.56 (-6.68, -6.44) | |
| 50 – 60 years | -10.13 (-10.32, -9.95) | <0.001 | -10.89 (-11.11, -10.68) | <0.001 | -7.25 (-7.40, -7.10) | <0.001 |
| <50 years | -9.61 (-9.83, -9.38) | | -10.88 (-11.15, -10.61) | | -7.17 (-7.36, -6.99) | |
| *Sex* | | | | | | |
| Female | -11.44 (-11.60, -11.29) | <0.001 | -11.25 (-11.43, -11.08) | <0.001 | -8.24 (-8.37, -8.11) | <0.001 |
| Male | -8.45 (-8.59, -8.31) | | -9.78 (-9.94, -9.62) | | -6.01 (-6.11, -5.91) | |

[1] Findings are reported as the difference in comparison to the lowest quintile. Negative values mean that the outcome of interest is smaller compared to the reference group (set as zero), while positive values indicate the opposite.

[2] P-values generated by including an interaction term between linoleic acid and age or sex independently in separate models.

**Supplementary Table 8: Longitudinal analyses between linoleic acid (% total fatty acid) expressed as per inter-quintile range and waist circumference, weight, and whole-body fat mass stratified by age and sex**

| | Waist circumference (cm) [1] | p-value [2] | Weight (kg) [1] | p-value [2] | Whole-body fat mass (kg) [1] | p-value [2] |
|---|---|---|---|---|---|---|
| *Age* | | | | | | |
| >60 years | -0.10 (-0.13, -0.06) | | -0.03 (-0.05, 0.00) | | -0.03 (-0.06, -0.01) | |
| 50 – 60 years | -0.10 (-0.13, -0.07) | 0.752 | -0.04 (-0.06, -0.02) | 0.022 | -0.04 (-0.07, -0.02) | 0.878 |
| <50 years | -0.09 (-0.13, -0.06) | | -0.04 (-0.07, 0.00) | | -0.03 (-0.07, 0.00) | |
| *Sex* | | | | | | |
| Female | -0.16 (-0.19, -0.13) | <0.001 | -0.06 (-0.08, -0.04) | 0.148 | -0.06 (-0.08, -0.03) | 0.546 |
| Male | -0.05 (-0.07, -0.02) | | -0.01 (-0.03, 0.01) | | -0.02 (-0.04, 0.00) | |

[1] Findings are reported as the difference in comparison to the lowest quintile in terms of change per year. Negative values mean that the outcome of interest is smaller compared to the reference group (set as zero), while positive values indicate the opposite.

[2] P-values generated by including an interaction term between linoleic acid and age or sex independently in separate models.
